# Nasopharyngeal *Staphylococcus aureus* colonization among HIV-infected children in Addis Ababa, Ethiopia: Antimicrobial susceptibility pattern and association with *Streptococcus pneumoniae* colonization

**DOI:** 10.1101/2022.11.24.22282703

**Authors:** Henok Alemu Gebre, Ashenafi Alemu Wami, Eyerusalem Solomon Kebede, Melaku Yidnekachew, Meseret Gebre, Abel Abera Negash

**Affiliations:** Armauer Hansen Research Institute (AHRI), Addis Ababa, Ethiopia

**Keywords:** *Staphylococcus aureus*, *Streptococcus pneumoniae*, MRSA, nasopharyngeal colonization, HIV-infected, pediatrics, Ethiopia, antimicrobial susceptibility

## Abstract

**Background:** *Staphylococcus aureus* and *Streptococcus pneumoniae* are common inhabitants of the nasopharynx of children. HIV-infected children have higher risk of invasive diseases caused by these pathogens. With widespread use of pneumococcal conjugate vaccines and the emergence of methicillin-resistant *S aureus*, the interaction between S. *aureus* and *S. pneumoniae* is of a particular significance. We sought to determine the magnitude of colonization by methicillin-sensitive and -resistant S. *aureus* and colonization by *S. pneumoniae*; associated risk factors and antimicrobial susceptibility pattern among HIV-Infected children in Addis Ababa, Ethiopia.

**Method:** A prospective observational study was conducted in 183 HIV-infected children at ALERT hospital Addis Ababa, Ethiopia from September 2016 to August 2018. S. *aureus* and *S. pneumoniae* were identified using standard bacteriological techniques, antimicrobial susceptibility testing was performed on S. *aureus* and screening for methicillin resistance was carried out by amplifying the *mecA* gene. Risk factors were analyzed by using binary logistic regression.

**Results:** The prevalence of nasopharyngeal *S. aureus*, MRSA and *S. pneumoniae* colonization were 27.3%, 2.7% and 43.2% respectively. Multivariable analysis indicated an inverse association between *S. aureus* and S. *pneumoniae* nasopharyngeal colonization (aOR, 0.49; CI, (0.24, 0.99); *p= 0*.*046*). The highest level of resistance in both methicillin sensitive *S. aureus* (MSSA) and MRSA was observed against tetracycline.

**Conclusions:** We found an inverse association between *S. aureus* and *S. pneumoniae* colonization among HIV-infected children. Continued assessment of the impact of pneumococcal conjugate vaccines and anti-retroviral therapy on nasopharyngeal bacterial ecology is warranted.

## Introduction

*Staphylococcus aureus* is a human commensal and also causes clinically important community and hospital acquired infections such as endocarditis, bacteremia, osteomyelitis and skin and soft tissue infections [1]. *S. aureus* colonizes multiple body sites in humans including anterior nares, skin, perineum, and pharynx. However, the most frequent carriage site for *S. aureus*, with a 27% carriage rate among adults is the nose and a causal relationship exists between *S. aureus* colonization and infection [2]. Compared to adults, persistent carriage is mostly seen in children and carriage rates vary between 45% in the first eight weeks and decline to 21% by six months of age [3].

Methicillin resistant *Staphylococcus aureus* (MRSA) was discovered in the 1960 in clinical isolates and then in the 1990s in the community. It has since spread globally and its prevalence is increasing resulting increased health care costs, morbidity and mortality [1,4]. HIV-infected patients, due to their weakened immune system, are at increased risk for several infections, including those caused by *S. aureus* and MRSA [5]. The morbidity and mortality associated with MRSA in patients with HIV infection [6] makes the study of MRSA colonization among the HIV-infected patients of particular interest. In addition, colonization with MRSA has been associated with 4-fould increase in the risk of infection [7].

Studies indicate that compared to healthy individuals, the prevalence of MRSA nasal colonization is higher in HIV-positive people [8,9].The risk of MRSA colonization often mirrors that of infection and additional people with higher risk of colonization/infection are, children, those in prisons, military recruits, those in poor neighbourhoods, livestock workers, individuals with prior MRSA infection and hospitalization and those with cystic fibrosis [1]. The main risk factors for colonization and infection among HIV-infected patients on the other hand are use of antimicrobials, previous hospitalization and low CD4+ T lymphocyte counts [5].

MRSA is now considered an urgent threat to public health and among the priority list of pathogens for which new antibiotics are required [10]. In order to make effective treatment of patients with disease due to MRSA and to prevent further transmission, accurate detection of methicillin resistance in is of the utmost importance [11].

The *mecA* gene is highly conserved in staphylococcal strains and thus is a useful marker and is considered as the gold standard for identifying methicillin resistance in *S. aureus* isolates. The *mecA* gene is located on the staphylococcal chromosome cassette *mec* and encodes penicillin binding protein 2a (PBP2a) [11]. Other chromosomal factors such as *femA* and *femB* are also associated with the expression of methicillin resistance[12].

Pneumococcal conjugate vaccines began being introduced in infant immunization schedules from the year 2000 and studies indicate that administration of PCVs to infants modifies not only the carriage of *Streptococcus pneumoniae* but also that of *S. aureus* and there is an inverse relationship between the carriage of the two bacteria [13,14]. There was also an earlier clinical trial that indicated an increase in acute otitis media due to *S. aureus* after PCV vaccination [15]. We have recently been able to show that 5-6 years after introduction of PCV10 in Ethiopia, *S. aureus* was the main cause bacteremic community acquired pneumonia among children [16].

Earlier studies in HIV infected children indicated a lack of association between *S. pneumoniae* and *S. aureus* carriage and cite suboptimal adaptive immunity as a possible reason [13] but there are suggestions that *S. pneumoniae – S. aureus* interference is CD4+ T cell-mediated and returns to normal following anti-retroviral therapy (ART) [17].

In Ethiopia, PCV10 was introduced in October 2011 as a three dose primary series (3p+0) without any booster dose and a decision has now been made to switch to PCV13. There is a scarcity of data in Ethiopia on the relationship between the nasopharyngeal carriage of *S. pneumoniae* and *S. aureus* and the possible impact of PCV introduction specialy among HIV-infected children. The aim of this study was therefore to determine the magnitude of methicillin-sensitive and -resistant *S. aureus* nasopharyngeal colonization and; their association with *S. pneumoniae* colonization among HIV-infected children in Addis Ababa, Ethiopia.

## Materials and Methods

### Study design and setting

A prospective observational study was conducted at All Africa Leprosy Rehabilitation and Training Hospital (ALERT), a governmental referral hospital, located in Addis Ababa, Ethiopia. The target population of this study was HIV-positive children aged 0-15 years coming for ART follow-up to the ALERT pediatric HIV clinic from September 1, 2016 to August 31, 2018.

### Sample Size

There is a population of approximately 750 HIV positive children who are provided HIV services at the ALERT pediatric ART clinic. We used an expected prevalence of nasopharngeal *S. auerus* colonization due to absence of studies that report prevalences among HIV-infected children in Africa in the post-PCV period. Accordingly, using a 95% confidence level, expected prevalence of 19.4% from a previous study in Uganda [18], 5% margin of error and design effect of 1, the minimum sample size required was 182.

### Patient enrollment and data collection

Trained research nurses approached the parents or guardians of the HIV-infected children who came for an initiation or followup visit at the ART clinic and briefed them on the information that has been provided in the patient information sheet. Children of parents who gave consent were then included in the study. Study participants’ demographic and clinical data including information on risk factors for colonization and disease were obtained and transferred to the questionnaire prepared for the study.

The dependent variable assessed was colonization with *S. aureus*. The independent variables assessed were: gender, age, number of household members, no of rooms in the house, presence of siblings <5 years of age, parents level of education, monthly income, exposure to cigarette smoke, reasons for hospital visit, antiretroviral treatment status, PCV vaccination, *S. pneumoniae* colonization, upper respiratory tract infection, lower respiratory tract infection within the last three months, CD4+ T cell count, and previous hospitalization within the last three months.

### Specimen collection

Nasopharyngeal (NP) swabs were collected from 183 HIV-infected consecutive children using flocked mini tip rayon swabs (Copan, Brescia, Italy). Briefly, tipping the head of the children tipped slightly backward, the swab was directly inserted parallel to the base of the NP passage until reaching the nasopharynx which is located about one-half to two-thirds the distance from the nostril to the ear lobe. Once the swab was fully passed into the nasopharynx, it was rotated 180_o_ and left in place for 5s to saturate the swab tip. The swab was then removed slowly and immediately transferred into cryovials containing 1 ml of STGG (skim-milk, tryptone, glucose, and glycerol) transport medium. The inoculated STGG was initially stored at -20°C in the hospital ward. Samples were then transported to the Armauer Hansen Research Institute (AHRI) laboratory using ice and were frozen at -80°C until further analysis.

### Culture and identification

The samples were initially thawed and 100-μl of the NP swab sample was plated on a blood agar plate (BAP) with and without gentamicin by streaking the sample using a sterile loop. BAP plates were then incubated at 37_o_C with ∼5% CO2 for 18-24 hours. *S. aureus* was identified using colony morphology on sheep blood agar, gram staining, catalase activity, production of coagulase, and growth on mannitol salt agar. Identification of *S. pneumoniae* was made on the basis of colony morphology, optochin susceptibility, and bile solubility.

### Antimicrobial susceptibility testing

The susceptibility of *S. aureus* isolates to chloramphenicol (30μg), ciprofloxacin (5 μg), clindamycin (2 μg), erythromycin (15 μg), gentamicin (10μg), tetracycline (30 μg), and trimethoprim/sulfamethoxazole (1.25 μg/23.75 μg) was determined by the disk diffusion method on Mueller-Hinton agar (Oxoid, Basingstoke, England). Test results were interpreted according to the Clinical and Laboratory Standard Institute (CLSI) criteria. *S. aureus* (ATCC® 25923) was used as a quality control organism.

### Detection of mecA genes

DNA extraction was performed by boiling *S. aureus* pellets in 300 μl of TE buffer as described previously [19]. Detection of 310-bp fragment of *mecA* was performed using primer pairs: 5′-GTAGAAATGACTGAACGTCCGATAA-3′ and 5′ CCAATTCCACATTGTTTCGGTCTAA -3′ as described previously [20]. The reaction mixture contained 12.5μl of hot star master mix (Qiagen, <u>Hilden, Germany</u>), 0.5 μl each of the forward and reverse primers, 9 μl of molecular grade water and 2.5μl of the template with a final volume of 25 μl. Amplification was carried out with 40 cycles of initial heat activation at 95_o_C for 15 minutes, denaturation at 94_o_C for 30 seconds, followed by annealing at 52_o_C for 45 seconds, extension at 72 _o_C for 1 minute, and final extension at 72 _o_C for 10 minutes.The PCR products were analyzed by electrophoresis on a 2% agarose gel.

### Statistical analysis

Data were initially entered in ReDCap (Vanderbilt University, Nashville, Tennessee), exported into Excel and analyzed using PASW Statistics 20 software (SPSS Inc., Chicago, Illinois). Sociodemographic, environmental and clinical characteristics were analyzed using descriptive statistics. Bivariate analysis, using binary logistic regression, was initially performed in order to determine factors associated with *S. aureus* colonization. Multivariate logistic regression was then used to assess independent associations for variables significant at *p* < 0.1. Variables at *p* < 0.05 were then considered statistically significant in the multivariable analysis.

### Ethical considerations

The study procedures were in accordance with the Helsinki Declaration. The study protocol was approved by the AHRI/ All Africa Leprosy Rehabilitation and Training Hospital (ALERT) Ethical Review Committee (AAERC) (PO/017/2015). Official permission letter was obtained from ALERT hospital. A written informed assent and consent was obtained from study participants, and parents or guardians of children respectively, before including them in the study. The study participant’s right to refuse or not give nasopharyngeal samples without affecting their routine medical services was granted. Samples were coded to keep the confidentiality of the study participants’ personal information. Authors access to information that could identify individual participants during or after data collection.

## Results

### Socio-demographic and clinical characteristics of the study participants

Out of potentially eligible 750 HIV-infected children, nasopharyngeal samples were collected from a total of 183 HIV-infected children at ALERT hospital. Among the study participants, 50.8% (93/183) were girls; the mean age of the study participants was 10.78 ± 2.68 years. The mean CD4+ T cell count of the study participants was 886.25 cells/mm_3_ and 95.6% (175/183) of the children have received full dose of PCV10 (Table 1).

**Table 1:** Socio-demographic, environmental and clinical characteristics and their association with *S. aureus* colonization among HIV-infected children aged <15 years at ALERT hospital, Addis Ababa, Ethiopia.

| Variable | Category | All children<br>( n=183) | Colonized with<br>*S. aureus* | Bivariate analysis |  |
| --- | --- | --- | --- | --- | --- |
|  |  | No. (%) | No. (%) | COR (95% CI) | P<sup>c</sup> |
| Gender | Male | 90(49.2) | 25(24.6) | 1.05 (0.55-2.01) | 0.892 |
|  | Female | 93(50.8) | 25(25.4) | Ref |  |
| Age category (in years) | 0-4 | 3(1.6) | 2(<1) | 5.18 (0.45-58.75) | 1.86 |
|  | 5-9 | 51(27.8) | 12(13.9) | 0.8 (0.37,1.69) | .550 |
|  | 10-14 | 129(70.4) | 36(35.2) | Ref |  |
| Number of<br>household members | <=3 | 37(20.2) | 11(10.1) | 0.65 (0.24-1.76) | .396 |
|  | 4-6 | 113(61.7) | 26(30.9) | 0.41 (0.17-0.97) | .065 |
|  | >=7 | 33(18) | 13(9.0) | Ref |  |
| Number of rooms in the<br>house | 1 | 69(37.7) | 16(18.9) | 0.71 (0.36-1.41) | 0.330 |
|  | >=2 | 114(62.3) | 34(31.1) | Ref |  |
| Siblings aged<5y | Yes | 68(37.2) | 13(18.6) | 0.5 (0.24-1.03) | 0.58 |
|  | No | 115(62.8) | 37(31.4) | Ref |  |
| Mother level of education | Illiterate | 68(37.2) | 22(18.5) | 2.1 (0.18-24.59) | 0.555 |
|  | First level | 72(39.3) | 18(19.7) | 1.25 (0.12-13) | 0.852 |
|  | Secondary | 39(21.3) | 9(10.7) | 1 (0.09-10.23) | 1.000 |
|  | College &above | 4(2.2) | 1(1.1) | Ref |  |
| **Father level of education** | Illiterate | 60(32.8) | 19(16.4) | 1.1 (0.26-5.03) | 0.860 |
|  | First level | 58(31.7) | 13(15.8) | 0.82 (0.2-3.27) | 0.773 |
|  | Secondary | 53(29) | 14(14.5) | 0.58 (0.15-2.23) | 0.426 |
|  | College & above | 12(6.6) | 4(3.3) | Ref |  |
| **Monthly income (in Ethiopian Birr)** | <1000 | 108(59) | 31(29.5) | 1.61 (0.43-6.10) | 0.483 |
|  | 1000-2500 | 43(23.5) | 11(11.7) | 1.38 (0.33-5.8) | 0.664 |
|  | 2500-3500 | 17(9.3) | 5(46) | 1.67 (0.32-8.6) | 0.541 |
|  | >3500 | 15(8.2) | 3(4.1) | Ref |  |
| **Exposure cigar ate smoking** | yes | 7(3.8) | 1(2.2) | 0.37 (0.04-3.1) | 0.355 |
|  | No | 176(96.2) | 49(47.8) | Ref |  |
| **Reason for hospital visit** | Asthma | 1(.5) | 0(0) | - | 1.000 |
|  | Dermatological problems | 18(9.8) | 5(4.9) | - | 1.000 |
|  | LRTI symptoms | 72(39.3) | 18(19.7) | 0.8 (0.42-1.61) | 0.570 |
|  | UTI | 7(3.8) | 3(1.9) | Ref |  |
| **Antiretroviral treatment status** | Follow-up | 149(81.4) | 41(40.4) | 0.9 (0.39-2.09) | 0.812 |
|  | Initiation | 34(18.6) | 9(9.6) | Ref |  |
| **PCV10 vaccine status** | Full dose<sup>a</sup> | 175(95.6) | 45(90) | 3.58 (0.92-13.93) | 0.065 |
|  | Incomplete<sup>b</sup> | 8(4.4) | 5(10) | Ref |  |
| ***S. pneumoniae* carriage** | Yes | 79(43.2) | 15(21.6) | 0.46 (0.23-0.93) | **0.029** |
|  | No | 104(56.8) | 35(28.4) | Ref |  |
| **URTI in the last 3 months** | Yes | 25(13.7) | 8(6.8) | 1.81 (0.61-5.4) | 0.495 |
|  | No | 158(86.3) | 39(41.8) | Ref |  |
| **LRTI in the last 3 months** | Yes | 6(3.3) | 2(1.6) | 1.34 (0.24-7.58) | 0.738 |
|  | No | 177(96.7) | 48(48.4) | Ref |  |
| **CD4<sup>+</sup> T cell count**<br>**( cells/mm3)** | >350 | 164(89.6) | 46(45.1) | 0.73 (0.23-2.34) | 0.600 |
|  | <350 | 19(10.4) | 4(4.9) | Ref |  |
| **Previous hospitalization in**<br>**the last 3 months** | Yes | 6(3.2) | 47(48.4) | 2.77 (0.54-14.18) | 0.223 |
|  | No | 177(96.7) | 3(1.6) | Ref |  |

**Legend:** COR: Crude odds ratio; CI: 95% confidence interval; CAP, community-acquired pneumonia; LRTI, lower respiratory tract infection; PCV, pneumococcal conjugate vaccine; URTI, upper respiratory tract infection; UTI, urinary tract infection; \*LRTI symptoms represent (cough, difficulty in breathing, and runny nose); <sup>a</sup>Full dose PCV10 indicates all three doses; <sup>b</sup>Incomplete PCV10 vaccination includes $\leq 2$ doses; <sup>c</sup>P value of binary logistic regression analysis, Values in bold: $P < 0.05$ .

### Prevalence of S. *aureus* and *S. pneumoniae* colonization

The overall prevalence of *S. aureus* nasopharyngeal colonization among the HIV-infected children in this study was 27.3% (50/183) whereas. *S. pneumoniae* was isolated from 43.3% (79/183) of the children (Table 1).

### Prevalence of MRSA colonization and detection of *mecA*

*mecA* was detected in 10% (5/50) of the *S. aureus* isolates and MRSA colonization rate was therefore 2.7% (5/183).

### Risk factors associated with *S. aureus* colonization

According to multivariable logistic regression analysis, there was a statistically significant association between *S. aureus* and *S. pneumoniae* nasopharyngeal colonization (aOR, 0.49; CI, (0.24-0.99); *p= 0*.*046*) (Table 2).

**Table 2:** Risk factors associated with *Staphylococcus aureus* colonization among HIV-infected children aged <15 years at ALERT hospital, Addis Ababa, Ethiopia.

| Variable | Bivariate |  | Multivariable |  |
| --- | --- | --- | --- | --- |
|  | cOR (CI) | p<sup>a</sup> | aOR (CI) | p<sup>a</sup> |
| PCV10 vaccine status | 3.58 (0.92-13.93) | 0.065 | 2.81 (0.7-11.34) | 0.148 |
| *S. pneumoniae* carriage | 0.46 (0.23-0.93) | **0.029** | 0.49 (0.24-0.99) | **0.046** |
| Number of household members (4-6) | 0.46 (0.2-1.05) | 0.065 | 0.49 (0.21, 1.15) | 0.104 |

**Legend:** PCV, pneumococcal conjugate vaccine; aOR, adjusted odds ratio; CI, 95% confidence interval.
<sup>a</sup>P value of binary logistic regression analysis. Values in bold: $P < 0.05$ .

### Antibiotic susceptibility pattern of S. *aureus* isolates

Antimicrobial susceptibility patterns were determined for all the 50 *S. aureus* isolates. The highest level of resistance was seen against tetracycline (32%) followed by erythromycin (24%) and trimethoprim-sulfamethoxazole (24%) (Fig1).

**Figure 1:**
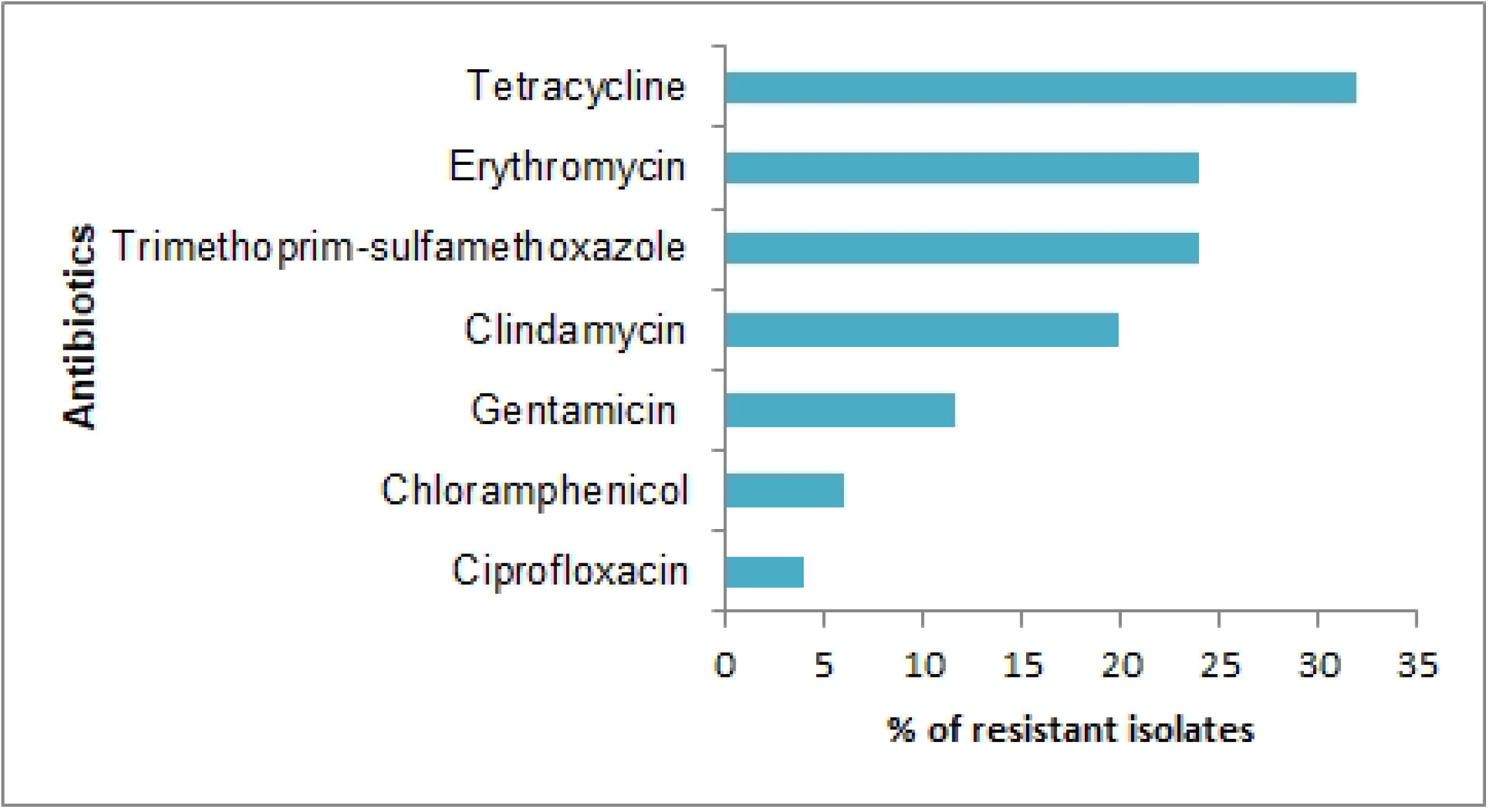
Antimicrobial susceptibility pattern of of *S. aureus* isolates among HIV – infected children aged <15 years at ALERT hospital, Addis Ababa, Ethiopia.

Among all MRSA isolates the highest percentage of drug resistance was seen against tetracycline (35.2%), erythromycin (29.4%), Clindamycin (29.4%), and gentamicin (11.7%), sulfamethoxazole-trimethoprim (5.9%), chloramphenicol (5.9%) and ciprofloxacin (5.9%). The overall multidrug-resistance (MDR) rate in this study was 12%.

## Discussion

Nasopharyngeal colonization and infection with *S. aureus* and MRSA is higher in HIV-infected children than those without HIV [21,22]. Determining the of S. aureus carriage rate and antibiotic resistance profiles is important in order to identify risk factors associated with *S. aureus infection*.

In the current study, we found that 5-7 years after introduction of PCV10 in Ethiopia, prevalence of *S. aureus* nasal carriage was 27.3% (50/183). Our result is in agreement with a similar study done in Northern Ethiopia (29%) [23]. Similar prevalence has also been reported from South Africa, prior to the introduction of PCVs (25.6%) [24]. Our finding was however lower than a report from results from a study in Brazil (45.16%) before the introduction of PCVs [22]. This might be because of geographical differences, sample size, the use of antiretroviral drugs and the introduction of PCVs.

Our finding indicates that the prevalence of MRSA nasopharyngeal colonization in HIV-infected children was 2.7% (5/183). Our finding is similar to nasal MRSA colonization reported in among children and adults in Northern Ethiopia (2.4%) [25]. Our findings were however lower than the pharyngeal colonization reported in a similar group of HIV-infected children in Northern Ethiopia (9.7%) [23]. In the study by Mulu *et al* and colleagues, methicillin resistance was determined using cefoxitin discs whereas in the current study, we used *mecA* amplification to determine methicillin resistance. Although Cefoxitin resistance is known as a reliable surrogate marker for mecA mediated methicillin resistance [26], there are studies that have questioned its accuracy [27]. Additional reasons for discrepancies in prevalence of MRSA could be differences study locations and periods, differences in the implementation of infection prevention among the hospitals and rational antibiotic use practices in the two settings.

In multivariable analyses, there was an inverse association between *S. aureus* and S. *pneumoniae* nasopharyngeal colonization in this group of HIV-infected children. Accordingly HIV-infected children who are colonized with S. *pneumoniae* are less likely to be colonized with *S. aureus*. A negative association between carriage of S. aureus and vaccine type *S. pneumoniae* was first reported close to two decades ago by two studies from Israel [28] and the Netherlands [29] before the introduction of pneumococcal conjugate vaccines. Since then, other studies from different parts of the world have reported similar results [30,31] mainly among healthy or HIV-uninfected children. However, studies among HIV-infected children from South Africa indicated lack of association between the nasopharyngeal colonization of *S. aureus* and *S. pneumoniae* [13,24]. The possible reasons for the lack of competition between the two bacteria in HIV-uninfected children included reduced mucosal immunity and therefore decreased immunological pressure and increased exposure to respiratory pathogens [24]. Other authors however suggested that a secondary host-related mechanism associated mainly with CD4+ T cells might play an important role in the pathway of interaction between *S. pneumoniae* and *S. aureus* [32]. The studies from South Africa did not study the correlation between CD4+ T cell counts and bacterial colonization or interaction. In the current study, the mean CD4+ T cell count of the study participants was 886.25 cells/mm3.

According to the Ethiopian national guideline for HIV prevention, care and treatment, for all children living with HIV, antiretroviral treatment (ART) is given as early as possible regardless of their WHO clinical stages and CD4+ T cell counts/percentage [33]. Studies indicate that early HIV diagnosis and early ART reduce infant mortality and HIV progression [34]. In addition, in HIV-infected infants with early ART treatment CD4+ T cell counts stabilize at the highest levels possible [35]. We therefore hypothesize the inverse interaction between *S. aureus* and *S. pneumoniae* in the HIV-infected children in this study similar to previous reports HIV-uninfected children is due to the impact of ART treatment.

People living with HIV are at increased risk of acquisition and infection with drug resistant bacteria [6]. In this study, among both MSSA and MRSA isolates, the highest level of resistance was seen against tetracycline which was consistent with a similar study among HIV-infected children in Northern Ethiopia [36] although higher percentage of resistance to tetracycline among MRSA isolates (72%) compared to our study (35.2%) were reported. The low rates of resistance to most of the antibiotics tested observed in this study might suggest that most of the MSSA and MRSA were community acquired strains [37].

Our study had some limitations. Firstly, prevalence of *S. aureus* colonization might have been underestimated since we sampled only sampled nasopharyngeal swabs. Since our results originate from only one hospital, the results might be fully representative of children in the whole of Addis Ababa or Ethiopia. Because serotyping was not performed for the *S. pneumoniae* isolates, we could not determine the relationship between vaccine type and non-vaccine type *S. pneumoniae* colonization and *S. aureus* colonization. In addition, our focus was only on colonization and the study was not designed to identify disease caused by these pathogens.

## Conclusions

The results of this study suggest that five to seven years after introduction of PCV10 in Ethiopia and more than 12 years after introduction of ART in Ethiopia the nasopharyngeal colonization rate of HIV-infected children at ALERT hospital, Addis Ababa, Ethiopia was 27.3% and 10% of the *S. aureus* isolates were MRSA. In addition, there was an inverse relationship between the colonization of *S. aureus* and *S. pneumoniae*.

## Data Availability

All relevant data are within the manuscript and its Supporting Information files.

## Acknowledgments

We would like to express our sincere gratitude to all of the children and guardians who consented to participate in the study. We would also thank the physicians and nurses who were involved in clinical diagnosis and sample collection and the Armauer Hansen Research Institute.

